# Knowledge and information sources of Helicobacter pylori among Jordanian population: a cross-sectional study

**DOI:** 10.1101/2022.11.10.22282172

**Authors:** Nader Alaridah, Raba’a F. Jarrar, Rayan M. Joudeh, Mallak Aljarawen, Mohammad Jum’ah, Hasan Nassr, Raad Riad AlHmoud, Abdullah Allouzi, Eslam M. Wadi, Anas H. A. Abu-Humaidan

**Author notes:** **Corresponding Author:**, Nader Alaridah MD, PhD.

## Abstract

In 2017, the Jordanian Ministry of Health reported that gastric cancer was one of Jordan’s most commonly diagnosed cancers. Gastric cancer is often linked to Helicobacter pylori, one of the foremost risk factors. Despite the high prevalence of H. pylori in Jordan, no information is available regarding the general population’s awareness of the harmful effects of this pathogen. The study aims to assess the knowledge and the impact of the source of knowledge on H. pylori among the general population in Jordan. A cross-sectional study involving 933 participants was conducted between May and July of 2021. After meeting the inclusion criteria and consenting to participate in this study, participants completed the questionnaire. The questionnaire was conducted interview-based, covering the following sections: sociodemographic data and knowledge related to H. pylori infection. 63% of the participants had a high level of education, 70.5% got their information about H. pylori infection from non-medical sources, and 68.7% had a low level of knowledge. Working in the medical field and attaining information from medical resources showed a significant association with a high level of knowledge, on the other hand, having a history of self-reported or family member of H.pylori infection also showed a significant association with a high level of knowledge. The Mann-Whitney U test showed that the mean ranks of all knowledge items in the medical source group significantly exceed those of the non-medical source group (p-values < 0.05). In Jordan, the awareness of H. pylori was unsatisfying, similar to other countries. Nevertheless, misconceptions in knowledge about H. pylori were identified and further awareness must be spread and advocated. Close observation of the non-medical sources of information is essential for delivering to the general population the sufficient amount of knowledge needed.

## Introduction

Nobody would have imagined, when *Helicobacter pylori* was discovered 35 years ago, that a bacterium measured in microns would grow to become a global concern spotlighting the interest of many researchers to dig deep and fetch more about this intriguing organism from the aspects that will help humanity deal with, eradicate, and diminish the complications arising from such infection. ^1^ Gastric cancer ranked as the fifth most common cancer worldwide with one million new cases which were associated with *H. pylori* infection by 2020.^2^ Eastern Asia had the highest rates of gastric cancer worldwide among both genders.^3^ Unfortunately, the Hashemite Kingdom of Jordan has no updated statistical or epidemiological findings about gastric cancer. In 2017, the Ministry of Health reported that gastric cancer has ranked ninth among the most commonly diagnosed cancers with 3.3% of the total cancer cases in Jordan.^4^ Moreover, it was evident that gastric cancer is the seventh and tenth most frequent cancer among Jordanian males and females, respectively. Statistical information about gastric cancer in Jordan is deficient due to the lack of any national screening system established in the kingdom. In addition, such high prevalence is similarly displayed in other developing countries around the world. ^5^ Interestingly, one cross-sectional study found that approximately 88% of the Jordanian population tested positive for *H. pylori*.^6^ On the other hand, developed countries have shown lower prevalence than those developing countries.^7^ This can be linked and associated with various poor hygienic routines of facilities and infrastructural bases.^8^ Knowledge is one of the most critical factors associated with the high prevalence of *H. pylori* among the Jordanian population. Globally, knowledge and awareness are inadequate regarding *H. pylori* infection among the general population.^9^ Lamentably and to our knowledge, there is no published data about the level of knowledge of the Jordanian general population for *H. pylori* infection. The value of the knowledge acquired is influenced greatly by the sources of information used. There are various sources of information one can use to obtain related health information including doctors, healthcare professionals, TV, radios, books, the internet, and social media networks. One study conducted in Jordan showed that specialists were the most used source of information among the general population.^10^ On the other hand, one cross-sectional study done in Saudi Arabia showed that participants are immensely influenced by the information provided through social media platforms.^11^ Such findings indicate the need to assess the validity, accuracy, and adequacy of different information sources on the amount of knowledge the general population has. The primary aim of this study is to assess the level of knowledge and investigate the sociodemographic factors and their relationship with the level of knowledge. The secondary aim is to identify numerous scopes of information sources that will influence the level of knowledge about *H. pylori* among the Jordanian general population as well as identify the areas of improvement needed to reach the optimum understanding and sufficient knowledge about *H. pylori* infection.

## Materials and methods

### Sample recruitment and study setting

Descriptive cross-sectional study was carried out in Jordan University Hospital between May and July of 2021. Outpatient Visitors of Jordan University Hospital were encouraged to voluntarily take part in the study. Participants had to be at least 18 years old, able to converse verbally in Arabic, and willing to engage in the study to be enrolled. Those who did not match the inclusion requirements were excluded. Random sampling method was used to recruit the participants from outpatient clinic visitors in different clinics. medical students from 5^th^ and 6^th^ year were trained by research team to interview the volunteered visitors. Participants who agreed to participate in this study answered interview-based questions about their *H. pylori* knowledge and the source of the information used. The questionnaire took 8 minutes to complete. The estimated minimum sample size needed for the study is 385, this was calculated based on a 5% margin of error and 50% prevalence.

### Development of the survey

We designed a questionnaire based on a literature review ^12^ and other studies ^13-15^. As there is no validated tool available to assess community knowledge. The questionnaire was written in Arabic. After the survey was finished, it was analyzed and verified by a gastroenterologist. Following the pilot testing, certain changes were made to ensure that the questions were comprehensible. The internal consistency was evaluated, and Cronbach’s alpha of knowledge was calculated (0.83)

### Measurement tool

Data were gathered using an Arabic-based online questionnaire composed of the following sections: participants’ background characteristics and general knowledge. Sociodemographic characteristics were obtained including age, gender (male, female), marital status (married, unmarried), educational level (high education, low education), occupational status (medical field, non-medical field, not working), self-reported History of *H*.*pylori* infection(yes, no),family history of H.pylori (yes, no), current residence (urban/rural housing), and source of information acquired about *H. pylori* (medical source [i.e., a doctor, nurses, pharmacist etc, or from educational field], non-medical sources[i.e. from a family/friends or social media or TV/Radio, etc]).

There questionnaire had 43 questions about different topic about *H*.*Pylori*, the questions divided into seven main items to assess the level of knowledge about *H. pylori*, the items as following: 1) nature of *H. pylori*. 2) organs affected by *H. pylori*, 3) mode of transmission, 4) common signs and symptoms, 5) methods of diagnosis, 6) type of treatment, 7) general knowledge about *H. pylori* including prevalence in Jordan, important risk factors, and complications. The answers for the questions were as the following: (Yes, no, I don’t know). Those who answered the question correctly were given 1 point who answered I don’t know 0 point were given. In this study, a cut-off value of 70% was used to categorize participants’ scores knowledge into a binary category (high knowledge and low knowledge). The questionnaire and the correct answers and its percentages for each question displayed in the **supplementary material file 1 and 2**.

### Ethical considerations

The Institutional Review Board (IRB) of the University of Jordan, Amman, Hashemite Kingdom of Jordan, reviewed and approved the study protocol in meeting No 2021/18 (reference number: 10/2021/27975) It was written in accordance with the Helsinki Declaration’s ideals. All participants were asked for their informed consent prior to the introduction of the questionnaire. Once the data was acquired, it was kept in absolute confidence and stored away, with only the lead investigator having access to it.

### Statistical analysis

Data were entered into Microsoft Excel (2016) and then imported into IBM SPSS version 25 (IBM Corp., Armonk, N.Y., USA) for analysis. For each quantitative and categorical variable, descriptive statistics were computed and expressed as either frequency and percentage or mean and standard deviation (SD). Associations between demographic variables and knowledge (binary level) were analyzed using the Chi-square test. Significantly associated variables were included in multivariate regression analysis controlled for potential confounders to assess the independent effect of each variable. Mann-Whitney U-test was used to evaluate the association between the source of information variable and the mean scores of the knowledge score items. A p-value of α < 0.05 and a Confidence Interval of 95% were set to determine the statistical significance of the reported results.

## Results

### Sociodemographic characteristics of the participants

A total of 933 participants met the inclusion criteria and completed the questionnaire. Their sociodemographic characteristics are demonstrated in **Table 1**. A number of 588 (63%) had a high educational level (had a diploma degree or higher). In addition, 119 (12.5 %) of the participants work in the medical field, Moreover, 165 (17.7%) of participants have a self-reported history of *H*.*pylori* infection and 301 (32.3%) of participants had a family history of *H*.*pylori*. Furthermore, 275 (29.5%) of the participants acquired information about *H. pylori* infection from medical sources.

**Table 1:** Sociodemographic characteristics of *H. pylori* questionnaire participants (n=933)

| Characteristics | (n/mean) | (%/SD) |
| --- | --- | --- |
| **Age** | 40.4 | ±16.1 |
| **Gender** |  |  |
| Male | 493 | 52.8 % |
| Female | 440 | 47.2 % |
| **Education Level** |  |  |
| Low Education | 345 | 37% |
| High Education | 588 | 63% |
| **Marital status** |  |  |
| Married | 602 | 64.5% |
| Unmarried | 331 | 35.5% |
| **Job Field** |  |  |
| Medical | 119 | 12.5% |
| Non-medical | 487 | 52.2% |
| No work | 327 | 35% |
| **Type of Residence** |  |  |
| City | 824 | 88.3% |
| Country | 109 | 11.7% |
| **Self-reported History of H.pylori infection** |  |  |
| Yes | 165 | 17.7% |
| No | 768 | 82.3% |
| **Family History of H.pylori** |  |  |
| Yes | 301 | 32.3% |
| No | 632 | 67.7% |
| **Source of information** |  |  |
| Medical | 275 | 29.5% |
| Non- medical | 658 | 70.5% |

### Knowledge of H. pylori and its associated factor

The summary of the participant’s correct answers is shown in **Supplementary Material file 2**. Out of the 933 participants, 292 (31.3%) and 641 (68.7%) had high and low knowledge levels, respectively. Regarding the nature of *H. pylori*, only (37.3%) stated that *H. pylori* belong to bacterial species. (76.7%) of the participants chose the stomach as the organ which *H. pylori* colonize. The participants were asked about the main route of *H. pylori* transmission and the percentage of correct answers were as the following: food (68.6), water (63.8%), and saliva (38.4%). For the signs and symptoms of *H. pylori*, the percentage of the correct symptoms were abdominal pain (75.5%), nausea/vomiting (68.4%), and abdominal bloating (59.8%). Regarding the treatment, (72.7%) of the responses were triple therapy as the main way to eradicate *H. pylori* infection. When the participants were asked about the diagnosis of *H. pylori*, the responses for the correct answers were as follows: stool antigen (62.2%), endoscopy (62.4%), and urea breath test (13.8%). Regarding general knowledge questions, (44.1%) of the participants acknowledged that more than half of the Jordanian population might be infected with *H. pylori*. (69.1%) and (55.2%) participants claimed that *H. pylori* can be attributed to the development of gastric and duodenal ulcers, respectively. As well as (59.5%) of the participants confirmed that *H. pylori* infection can be a life-threatening condition if left untreated. (60.6%) and (63.2%) of the participants asserted that spicy food and chronic stress may worsen the symptoms of *H. pylori* infection, respectively. When participants were asked about the long-term risk of developing malignancy from chronic *H. pylori* infection, (45.7%) and (26.6%) of the participants responded that *H. pylori* infection can lead to gastric cancer and mucosa-associated lymphoid tissue (MALT) B-cell lymphomas, respectively.

Some sociodemographic variables were statistically significantly associated with knowledge as shown in **Table 2**. Multivariate logistic regression analysis showed significant association with the following variables: job field (Medical: OR=2.936; 95% CI= 1.718-5.018, p= 0.000; ref: No-work), educational level (Higher education: OR=1.948; 95% CI= 1.355-2.801, p= 0.000; ref: Lower educational level), self-reported history of H.pylori (Yes: OR=2.119; 95% CI= 1.392-3.225, p= 0.000; ref: No), family history of H.pylori (Yes: OR=2.129; 95% CI= 1.545-2.935, p= 0.000; ref: No), source of knowledge (medical source: OR=2.224; 95% CI: 1.535-3.221, p-value: 0.000; ref: non-medical source). Neither age, gender, marital status, nor residency was statistically associated with the knowledge regarding *H. pylori according to th*e multivariate logistic regression analysis, as reported in **Table 3**.

**Table 2:** Association between sociodemographic characteristics of the general population and knowledge about H. pylori (n= 933)

| *Variables* | *Level of Knowledge* |  | *P-value* |
| --- | --- | --- | --- |
|  | *High (n=292)* | *Low (n=641)* |  |
| *Age (mean/SD)* | 38.5(15±) | 41.2(16.5±) | 0.014 |
| *Gender:* |  |  |  |
| Female | 156 | 284 | 0.010 |
| Male | 136 | 357 |  |
| *Marital status:* |  |  |  |
| Married | 177 | 425 | 0.092 |
| Unmarried | 115 | 216 |  |
| *Education level:* |  |  |  |
| Lower education | 74 | 271 | 0.000 |
| Higher education | 218 | 370 |  |
| *Job field:* |  |  |  |
| Medical | 76 | 43 | 0.000 |
| Non-medical | 125 | 362 |  |
| No-work | 91 | 236 |  |
| **Residence:** |  |  |  |
| City | 261 | 559 | 0.494 |
| Village | 31 | 78 |  |
| **Self-reported History of H.pylori infection** |  |  |  |
| Yes | 83 | 82 | 0.000 |
| No | 209 | 559 |  |
| **Family History of H.pylori** |  |  |  |
| Yes | 123 | 178 | 0.000 |
| No | 169 | 463 |  |
| **Source of information:** |  |  |  |
| Medical | 148 | 127 | 0.000 |
| Non- medical | 144 | 514 |  |

**Table 3:** multivariate logistic regression associations between knowledge and the sociodemographic characteristics of the general population in Jordan.

| Covariates | Knowledge |  |  |
| --- | --- | --- | --- |
|  | OR | CI | p-value |
| Age 38.5(15±) | 0.996 | 0.986-1.006 | 0.349 |
| **Gender** |  |  |  |
| Male |  | Reference |  |
| Female | 1.348 | 0.998-1.918 | 0.051 |
| **Education level:** |  |  |  |
| Lower education |  | Reference |  |
| Higher education | 1.948 | 1.355-2.801 | 0.000 |
| **Job field:** |  |  |  |
| Medical | 2.936 | 1.718-5.018 | 0.000 |
| Non-medical | 0.852 | 0.554-1.161 | 0.411 |
| No-work |  | Reference |  |
| **Self-reported History of H.pylori infection** |  |  |  |
| Yes | 2.119 | 1.392-3.225 | 0.000 |
| No |  | Reference |  |

| Family History of H.pylori |  |  |  |
| --- | --- | --- | --- |
| Yes | 2.129 | 1.545-2.935 | 0.000 |
| No | Reference |  |  |
| *Source of information:* |  |  |  |
| Medical | 2.224 | 1.535-3.221 | 0.000 |
| Non- medical | Reference |  |  |

**Table 4:**
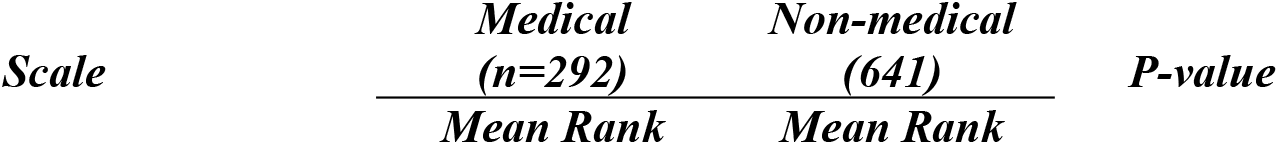
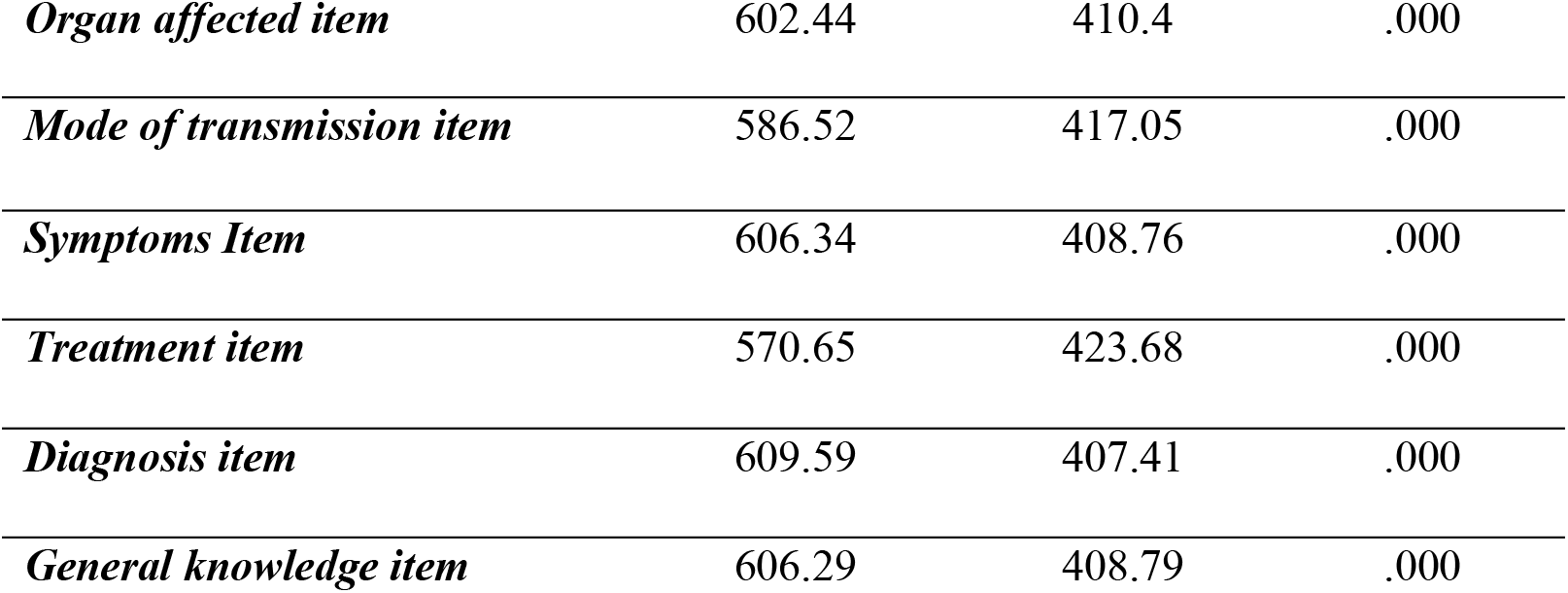
Mann-Whitney Test Analysis of Medical vs Non-medical Sources of Information regarding the questioned items.

### The sources of information and the level of knowledge acquired

As shown in **Table 1**, the participants in the study utilized various sources of information to attain their knowledge about *H. pylori* infection. The majority of the participant 658(70.5%) used non-medical sources (e.g., family, friends, social media, and TV) to obtain the information. In contrast, only a third 275 (29.5%) of the participant got their information about *H. pylori* infection, from medical sources such as from a doctor or during their study. By dividing the source of information about *H*.*pylori* infection into medical and non-medical sources; a significant statistical association between the source of information and the level of knowledge has been found (p-value = .000). The Mann-Whitney U test showed that the mean rank of all knowledge items in medical source category significantly exceeds those of the non-medical source group category (p-values < 0.05). The mean ranks and p-values of each question asked are shown in **Table 5**.

## Discussion

Few studies have been conducted to examine the general population’s awareness regarding *H. pylori* infection. Investigating public awareness of *H. pylori* infection might aid in determining various methods by which one’s understanding of the bacterium can be improved. Such improvements are expected to enhance the quality of life for many people as well as prevent the long-term consequences of chronic *H. pylori* infection. To the best of our knowledge, this study is the first of its nature in Jordan to report the level of knowledge about *H. pylori* infection and its predictors. It is also necessary to investigate the knowledge gap among the general population concerning the source of information.

This study included over 900 respondents from outpatient visitors of Jordanian university hospital. In general, the Jordanian population seems to have diminished awareness of *H. pylori* infection. Furthermore, the participant responses showed low awareness in the following items: The nature of *H. pylori*, Organ colonized by *H. pylori, H. pylori* signs and symptoms that are frequent, the definitive treatment and diagnosis and general knowledge about *H. pylori* prevalence in Jordan. As well as long-term *H. pylori* sequelae include the development of stomach cancer and mucosa-associated lymphoid tissue (MALT) B-cell lymphomas. In general, we discovered that respondents who obtained their knowledge from medical sources scored more than those who obtained their information from non-medical sources on all items.

Only one-third of the participants in our study reported to have high level of knowledge toward H.pylroi infection. This low percentage is consistent with the findings of previous research in our region.^13-15^ Notably, around half of the participants were aware that persistent *H. pylori* infection may lead to the development of gastric and duodenal ulcer diseases and gastric cancer. This study showed better findings compared to previous studies ^13^-^16^ and is quite similar to research done in Korea.^17^ Some sociodemographic characteristics were shown to be connected with greater levels of knowledge in this study. Females have much more information about *H. pylori* infection than males, according to our findings. This is in line with the findings of previous studies^13-15^ and might be linked to the fact that women are often having the role of the family caregiver, and so have more interactions with clinicians and the opportunity to learn about various diseases. Higher education level and working in the medical area, as predicted, have higher awareness *H. pylori* infection which has been documented in prior studies. ^13-15^ in our study, we found respondent who had history of *H*.*pylori* infection or had family members who diagnosed with *H*.*pylori* before, they tend to have high knowledge, this finding supported by the fact, upon diagnosing more information provided by the physician about *H*.*pylori*. in line with previous point, the authors found that obtaining information from medical sources was associated with a high level of knowledge.

The current study found unsatisfactory outcomes in terms of awareness. This is consistent with comparable research conducted in China, the United Arab Emirate and south Korea. ^13-15,17^. Misconceptions in critical areas were discovered. For example, more than half of the participants were not believing in any association between *H. pylori* infection and gastric cancer or Mucosa-associated lymphoid tissue (MALT) B-cell lymphomas. This stresses the importance of intervening to correct those misconceptions and to advocate more about how serious *H*.*pylroi* infection is. Many campaigns can be created to raise awareness among Jordanians through social media platforms and television. Moreover, we believe that health education can aid in the prevention of *H. pylori* infection. Thus, this will reduce its impact on the health care system and population. Health care providers should pay special attention to such dilemmas and implement curricular changes with an emphasis on preventive education. As shown in this study, the non-medical sources are the dominating sources used by the participants. Hence, we put forward and suggest intensive, close surveillance issued by the Jordanian Ministry of Health on social media platforms, TV, and radios to limit and control false and poorly evident health-related information. Thus, it will improve and enhance the knowledge of the population on gaining and obtaining information from authenticated and trusted sources such as healthcare providers.

## Strengths and limitations

To the best of our knowledge, this is the first survey that has evaluated the degree of knowledge among Jordanians. One of the study’s merits is the face-to-face manner performance, which allowed us to obtain a high response rate and reduce participation bias. This is the first research to look specifically at *H. pylori* awareness. The convenience sample approach was utilized, and single center involved, may have influenced the generalizability of the results. Self-reported data such as History of *H. pylori* infection among the participant and their family, makes the finding vulnerable to recall bias. In addition, the lack of a validated questionnaire is one of the barriers we encountered. The authors created this survey since there was no validated questionnaire available to test knowledge. Hence, a validated questionnaire must be developed in the future. Since this is a cross-sectional study, it is difficult to establish a cause-and-effect link. Furthermore, only quantitative measures were utilized to assess awareness. Indeed, sociocultural aspects such as beliefs and tradition were not investigated in our study; these factors should be investigated further in future studies.

## Conclusion

In conclusion, people in Jordan have low awareness regarding *H. pylori* infection. Having higher education degree, working in the medical field, history of *H*.*pylori*, family history of *H. pylori* and acquiring information from medical sources correlated with a high awareness of *H. pylori* infection. Interventions should be implanted to correct the flaws and misconceptions noted in the study to improve general awareness. Developing social programs and campaigns to raise awareness among the general population is a public health imperative in Jordan. Knowing the general principles of *H. pylori* infection will reduce the prevalence of *H. pylori* infection. In addition, encouraging the population to get information about *H. pylori* infection from trusted sources will enhance the understanding of the general population about routes of transmission and thus diminishes the consequences of *H. pylori* infection including the occurrences of gastric cancer secondary to such infection.

## Data Availability

All relevant data are within the manuscript and its Supporting Information files.

## Acknowledgment

We wish to express our gratitude to all administrative staff and student representatives from Jordanian medical schools for their facilitation and distribution of the survey. In addition, we would like to thank the nurses in the gastroenterology clinic for their help in data collection. Also, we would like to thank Dana Nedal Mohammad Rajab Abuaysheh for her contribution to organizing and grammatically reviewing the manuscript.

## Statements of ethics

This study was conducted ethically in accordance with the World Medical Association Declaration of Helsinki. Written informed consent was obtained from participants to participate in the study. The study’s protocol was ethically reviewed and approved by The Institutional Review Board (IRB) at the University of Jordan, Amman, the Hashemite Kingdom of Jordan (reference number: 10/2021/27975) in meeting No 2021/18.

## Author Contributions

NA supervised the team, designed the study concept and methodology RMJ and RFJ, performed most of the analysis, and was significantly involved in writing and revising the manuscript. RMJ and RFJ contributed to data collection, performed data analysis, and were highly involved in manuscript writing. MA, HN,AA2, and RA assisted with the development of the questionnaire, performed most of the literature review, and were heavily involved in the writing and editing of the manuscript. MJ, EMW, AA1 and MA assisted in data gathering, verified the accuracy of the results, and critically revised the manuscript. The final manuscript was reviewed and approved by all the authors.

### Data availability statement

Data are available uploaded on the supporting information.

